# Variants in *BSN*, encoding the presynaptic protein Bassoon, result in a novel neurodevelopmental disorder with a broad phenotypic range

**DOI:** 10.1101/2025.02.10.25321755

**Authors:** Stacy G Guzman, Sarah M Ruggiero, Shiva Ganesan, Colin A Ellis, Alicia G Harrison, Katie R Sullivan, Zornitza Stark, Natasha J Brown, Sajel L Kana, Anabelle Tuttle, Jair Tenorio, Pablo Lapunzina, Julián Nevado, Marie T McDonald, Courtney Jensen, Patricia G Wheeler, Lila Stange, Jennifer Morrison, Boris Keren, Solveig Heide, Meg W Keating, Kameryn M Butler, Mike A Lyons, Shailly Jain, Mehdi Yeganeh, Michelle L Thompson, Molly Schroeder, Hoanh Nguyen, Jorge Granadillo, Kari M Johnston, Chaya N Murali, Katie Bosanko, T Andrew Burrow, CHOP Birth Defects Biorepository, Penn Medicine Biobank, Syreeta Morgan, Deborah J Watson, Hakon Hakonarson, Ingo Helbig

## Abstract

Disease-causing variants in synaptic function genes are a common cause of neurodevelopmental disorders and epilepsy. Here, we describe 14 individuals with *de novo* disruptive variants in *BSN*, which encodes the presynaptic protein Bassoon. To expand the phenotypic spectrum, we identified 15 additional individuals with protein-truncating variants (PTVs) from large biobanks. Clinical features were standardized using the Human Phenotype Ontology (HPO) across all 29 individuals, which revealed common clinical characteristics including epilepsy (13/29 45%), febrile seizures (7/29 25%), generalized tonic-clonic seizures (5/29 17%), and focal onset seizures (3/29 10%). Behavioral phenotypes were present in almost half of all individuals (14/29 48%), which comprised ADHD (7/29 25%) and autistic behavior (5/29 17%). Additional common features included developmental delay (11/29 38%), obesity (10/29 34%), and delayed speech (8/29 28%). In adults with *BSN* PTVs, milder features were common, suggesting phenotypic variability including a range of individuals without obvious neurodevelopmental features (7/29 24%). To detect gene-specific signatures, we performed association analysis in a cohort of 14,895 individuals with neurodevelopmental disorders (NDDs). A total of 66 clinical features were associated with *BSN*, including febrile seizures (p=1.26e-06) and behavioral disinhibition (p = 3.39e-17). Furthermore, individuals carrying *BSN* variants were phenotypically more similar than expected by chance (p=0.00014), exceeding phenotypic relatedness in 179/256 NDD-related conditions. In summary, integrating information derived from community-based gene matching and large data repositories through computational phenotyping approaches, we identify *BSN* variants as the cause of a new class of synaptic disorder with a broad phenotypic range across the age spectrum.

## Introduction

Variants in genes linked to synaptic function have emerged as common contributors to neurodevelopmental disorders and epilepsy.^1,2^, At the presynaptic active zone, the protein encoded by the *BSN* gene (MIM: 604020) functions as a scaffolding protein that coordinates the positioning of synaptic vesicles and organizes molecular components critical for rapid neurotransmitter release, supporting precise synaptic signaling and plasticity.^3–5^ Disruption of such genes is a known causal mechanism for neurodevelopmental disorders, including those caused by variants in *SHANK3* (MIM: 606230), *SYNGAP1* (MIM: 603384), and *DLG4* (MIM: 602887).^6,7,8^ Disorders of synaptic function are increasingly associated with clinical phenotypes spanning epilepsy, autism spectrum disorder (ASD), and intellectual disability.^6,7,9,10^

*BSN* is highly expressed in the brain, and several *Bsn* deficient mouse models suggest its potential link to seizures.^11–14^ In *Bsn* mutant mice, the loss of functional BSN protein disrupts synaptic ribbon architecture in the retina and impairs presynaptic function, leading to sensory deficits and epileptic seizures. ^12,14–16^ Although *BSN* has been linked to brain disorders, few clinical cases with *BSN* variants have been reported, leaving the associated phenotypic spectrum unclear. Prior studies suggested variation in the *BSN* gene as a contributor to epilepsy with febrile seizures and a largely favorable outcome.^17^ However, the full spectrum of *BSN*-related phenotypes in larger cohorts, including the phenotypic consequences of *de novo* and inherited or unknown protein-truncating variants (PTVs), has not been assessed to date.

To further delineate *BSN*-related phenotypes, we leverage the Human Phenotype Ontology (HPO), a standardized framework that harmonizes clinical data across large, heterogeneous cohorts.^18–21^ By mapping phenotypic features to HPO terminology, subtle phenotypic patterns can be uncovered that might otherwise be obscured.^20,21^ Previous studies have demonstrated the power of using the HPO in large-scale genetic research, where it has been used to identify novel gene-phenotype associations.^19–23^ For example, *AP2M1* (MIM: 601024) was implicated in epilepsy and neurodevelopmental disorders through the characterization of individuals carrying *de novo* variants, highlighting the role of endocytosis in synaptic function.^22^ Accordingly, by systematically analyzing phenotypic similarities, HPO helps bridge the gap between genotype and phenotype, providing critical insights into the genetic basis of complex disorders.

Here, we applied the HPO framework to a cohort of 29 individuals with *BSN* variants, including 14 individuals with *de novo* variants, 13 individuals with PTVs of unknown inheritance, and 2 individuals with PTVs with paternal inheritance. Affected individuals presented with diverse neurodevelopmental phenotypes, including behavioral abnormalities, delayed speech, learning disabilities, and variable seizure types. By harmonizing phenotypic features through an HPO-based approach, we explored the phenotypic landscape of *BSN*-related disorders and examined the variability of phenotypes vary across the age span.

## Material and Methods

### Participant recruitment

We identified individuals with *BSN* variants through multiple sources, ensuring all variants were either *de novo* or protein-truncating (PTVs) or missense variants. Two participants were enrolled in the Epilepsy Genetics Research Project (EGRP, IRB 15–12226) cohort at Children’s Hospital of Philadelphia (CHOP). Both individuals had *de novo* PTV *BSN* variants identified through diagnostic trio whole exome sequencing (WES). Clinical data for these research participants were manually extracted from their electronic health records (EHR). An additional 14 individuals were identified through Genematcher^24^, an online platform that facilitates international collaborations by matching researchers and clinicians with overlapping genetic findings. Among these, nine individuals had confirmed *de novo BSN* variants (2 missense and 7 PTV), verified by their respective institutions.

Seven individuals with *BSN* PTVs were identified through the Penn Medicine BioBank (PMBB), a large-scale initiative that integrates genomic data with longitudinal EHRs. PMBB enrolls participants through in-person encounters at Penn Medicine outpatient sites, where written informed consent is obtained. All participants consented to the use of their de-identified data and test results for future research. The PMBB operates under IRB protocol #813913, with approval from the Institutional Review Board at the University of Pennsylvania.

One individual with a missense *de novo BSN* variant was identified from the Birth Defects Biorepository (BDB) at Children’s Hospital of Philadelphia. The BDB is an IRB-approved protocol (#18-015525), designed to store and permit access to biological specimens and longitudinal clinical and research data for future studies on birth defects. All participants consented to the use of their de-identified data and test results for future research on children with birth defects. Three additional individuals with *BSN* PTVs were identified through the Center for Applied Genomics (CAG) at Children’s Hospital of Philadelphia, a pediatric genomics research program focused on complex traits and rare diseases.

A literature review identified two previously reported individuals with *de novo* PTVs in *BSN*.^17^ Clinical data from the prior report was translated to HPO terms. While the exact ages of these individuals were not reported, the available clinical data were collected during infancy or early childhood, with seizure outcomes documented up to three years of age. Both individuals were included in the analysis of pediatric individuals with *BSN* variants.

### Variant identification and annotation

Variants identified through trio WES and were confirmed using standardized protocols as described previously.^22^ Diagnostic sequencing for individuals identified through GeneDx^25^ (Individuals 4, 6, 7, 9, 10) was conducted using exome capture platforms such as the IDT xGen Exome Research Panel v1.0 or v2.0 (Integrated DNA Technologies) and Twist Bioscience Exome 2.0 (Twist Biosciences), followed by massively parallel sequencing on Illumina platforms with paired-end reads of >100 bp. Sequencing data were aligned to the human genome reference build GRCh37/UCSC hg19, and variants were called using institution-specific pipelines, ensuring high-quality annotation and filtration.

For participants recruited through the EGRP cohort at CHOP, CAG, and PMBB, variant annotations were performed using ANNOVAR^26^. Additional variant filtration criteria included allele frequency (AF) < 0.005 (based on gnomAD v4)^27^ and pathogenicity predictors such as CADD > 15, REVEL > 0.2, and Genotype Quality (GQ) > 30. These thresholds ensured the retention of rare, likely pathogenic variants. Variants identified in Genematcher cohorts were confirmed by respective contributing institutions following previously published standards for variant interpretation. The sequencing and annotation methods used across all contributing cohorts were consistent with best practices for genomic analyses and align with protocols previously described in the literature.^22^

For the single individual identified through BDB, whole genome sequencing and data processing were performed by the Genomics Platform at the Broad Institute of MIT and Harvard. DNA libraries were prepared using the Illumina Nextera or Twist exome capture (∼38 Mb target) and sequenced with 150 bp paired-end reads, achieving >85% of targets covered at 20x and a mean target coverage of >55x. Sequencing data were processed through a pipeline based on Picard, with read mapping performed using the BWA aligner to the human genome build 38 (GRCh38). Variants were called using the Genome Analysis Toolkit (GATK) HaplotypeCaller package version 3.5, following best practices for variant detection.

### Phenotypic analysis

Clinical phenotypes from EGRP cohort at CHOP, PMBB, and CAG participants were confirmed through manual review of EMRs, ensuring accurate phenotypic information. All clinical data associated with the research participants were mapped to Human Phenotype Ontology (HPO) terms. For phenotyping forms and databases, we manually mapped clinical terms to HPO terms (HPO version 1.2; release format-version: 1.2; data-version: releases/2023-10-09; downloaded on 11/10/23) in accordance with prior studies.^23^ The phenotypes of all individuals were manually coded by expert reviewers. Phenotypes were first extracted by research staff with clinical and biomedical knowledge and experience with the HPO by using all available clinical and research notes for an individual and by using the most specific HPO terms applicable. These assigned terms were then reviewed and verified by domain experts, either a physician or genetic counselors specialized in epilepsy genetics. In cases of ambiguity and uncertainty, a higher-level, more general HPO term was coded rather than a more specific term.

For each individual, all higher-level (ancestral) HPO terms were derived, as previously reported.^22,23,28^ This method, known as propagation, results in a base and propagated set of HPO terms for each individual.^22,23,28^ The propagated HPO dataset from the entire cohort was used to generate baseline frequencies (f) for all HPO terms. Information content (IC) of each term was defined as the −log2(f), with a higher IC value reflecting a more specific and less frequently encountered HPO term in the cohort. In the current manuscript, we use a compact internationalized resource identifier (CURIE) to refer to HPO terms, i.e., “HP:0001250” (“Seizures”) abbreviates “https://hpo.jax.org/app/browse/term/HP:0001250” in accordance with the Open Biological and Biomedical Ontologies (OBO) Citation and Attribution Policy as previously described.^20,28^ For readability of the manuscript, we omit quotation marks for phenotypes expressed in HPO terms, streamline their descriptions, and adjust the grammatical usage of these terms within sentences. When followed by HPO identifier [e.g. “[…] seizures (HP:0001250)”], a phenotype refers to a clinical term coded in HPO terms rather than a more general reference to this phenotype.

For PMBB and CAG cohorts, ICD-9 and ICD-10-CM codes were provided as part of the datasets and were translated into HPO terms using a predefined mapping table.^29,30^ Longitudinal clinical data for these research participants were also provided and incorporated into the phenotypic analysis. Additionally, the ICD-to-HPO mapping process and longitudinal data integration underwent quality control steps to ensure robust phenotypic alignment across cohorts. This standardization facilitated a uniform phenotypic analysis across all cohorts.

### Data integration and processing

All datasets were curated to ensure consistency in variant annotation and phenotypic mapping. This process included manual validation steps to ensure accuracy in EHR data extraction, ICD-to-HPO mapping, variant filtration, and cohort selection.

### Statistical and computational analyses

All computations were performed using the R Statistical Framework. To assess the association between *BSN* variants and phenotypic features, we utilized statistical and computational methods aligned with those detailed in our previous publication.^28^ Volcano plots were generated to visualize association results, plotting –log_10_(p-value) against log_2_(odds ratio), and deriving p-values through Fisher’s exact tests.

Phenotypic similarity (sim) analyses were conducted using the simmax algorithm due to its established use in prior studies.^22,28^ Permutation testing (100,000 iterations) was employed to validate the statistical significance of phenotypic clustering, ensuring observed similarities exceeded those expected by chance. Specifically, the median similarity score for each gene was compared to a null distribution derived from random permutations of phenotypic data. The *denovolyzeR* tool was used to determine the probability of n *de novo* variants in a given gene.^31^

## Results

### Identification of two *de novo BSN* frameshift variants in individuals with early-onset seizures

We identified two individuals with novel *de novo* frameshift variants in the *BSN* gene through clinical exome sequencing. The *BSN* (NM_003458.4) variants in Individual #1 [c.8158_8162delACGGA (p.Thr2720Afs*38)] and Individual #2 [c.867_867dup (p.Pro290Afs*27)] were absent in the gnomAD (**Figure 1**). *BSN* is predicted to be highly intolerant to genomic variation that would lead to loss-of-function variation, with a probability of loss-of-funtion intolerance (pLI) score of 1.^27^

**Figure 1.**
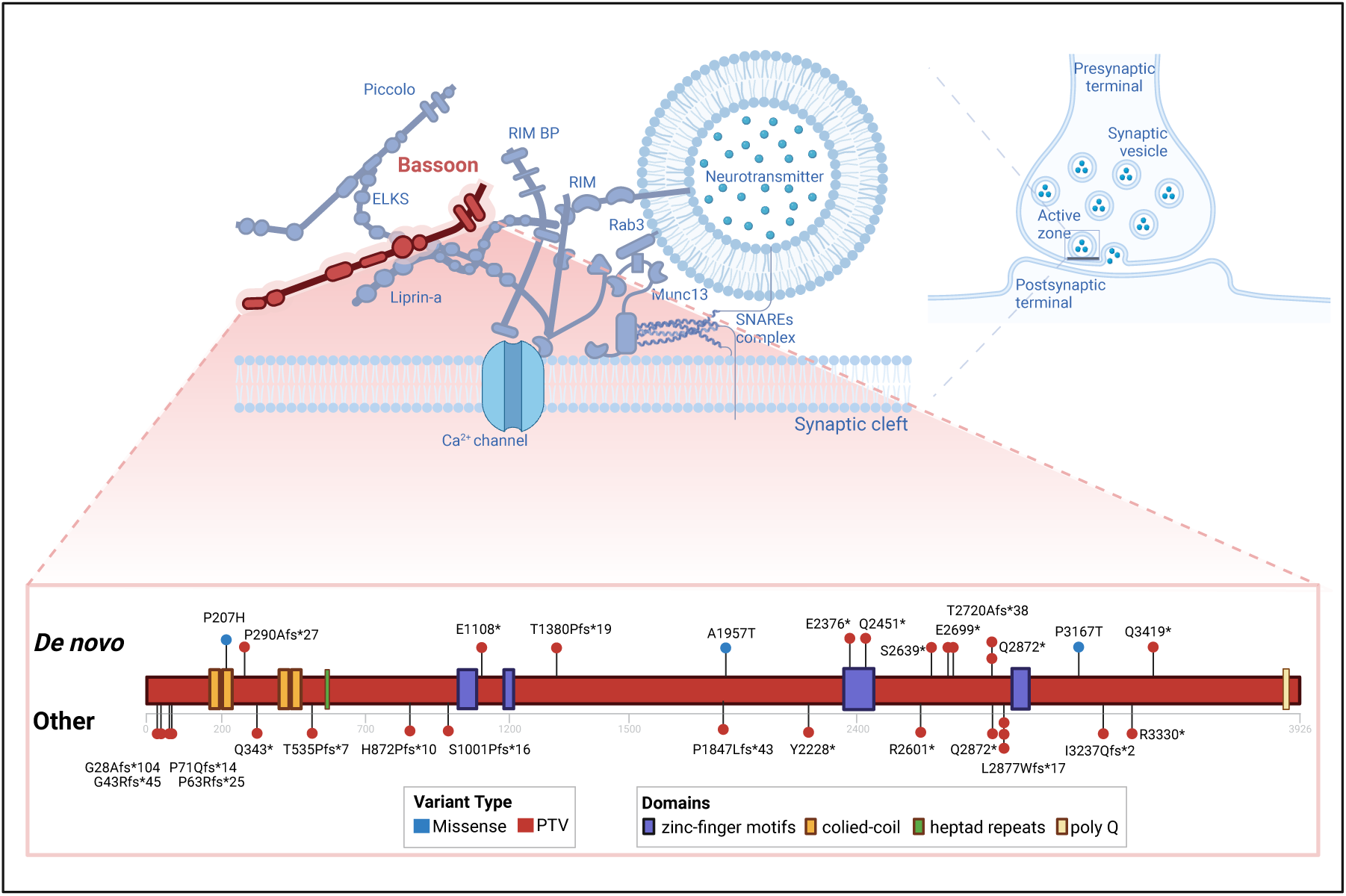
Overview of *BSN* Variants Identified in 29 individuals. (A) Diagram of presynaptic active zone assembly with synaptic vesicle fusion machinery proteins, showing BSN (red) as a key scaffolding protein in synaptic vesicle positioning and release. (B) *BSN* gene with variant distribution, where *de novo* variants (top) include both missense (blue) and protein-truncating variants (PTVs, red), while “Other” shows inherited and unknown inheritance variants (bottom) consist of PTVs only.

Both individuals presented with febrile seizures before 18 months of age (**Table 1**). They remained seizure-free until early childhood (range: 5–10 years), when Individual #1 had a first unprovoked bilateral tonic-clonic seizure, and Individual #2 presented with absence seizures. For Individual #1, seizures were infrequent initially and were managed with levetiracetam. By early adolescence (range: 10–15 years), Individual #1 started to have monthly bilateral tonic-clonic seizures, accompanied by a decline in academic performance. Individual #2 had infrequent absence seizures, followed by focal impaired awareness seizures and generalized tonic-clonic seizures. Both individuals exhibited behavioral abnormalities in early childhood and were diagnosed with ADHD different ages (one in early childhood, the other in late childhood). Individual #1 had early developmental delays, particularly in language, and was diagnosed with autism in early childhood. Both individuals had learning disabilities that necessitated specialized schooling.

**Table 1.**
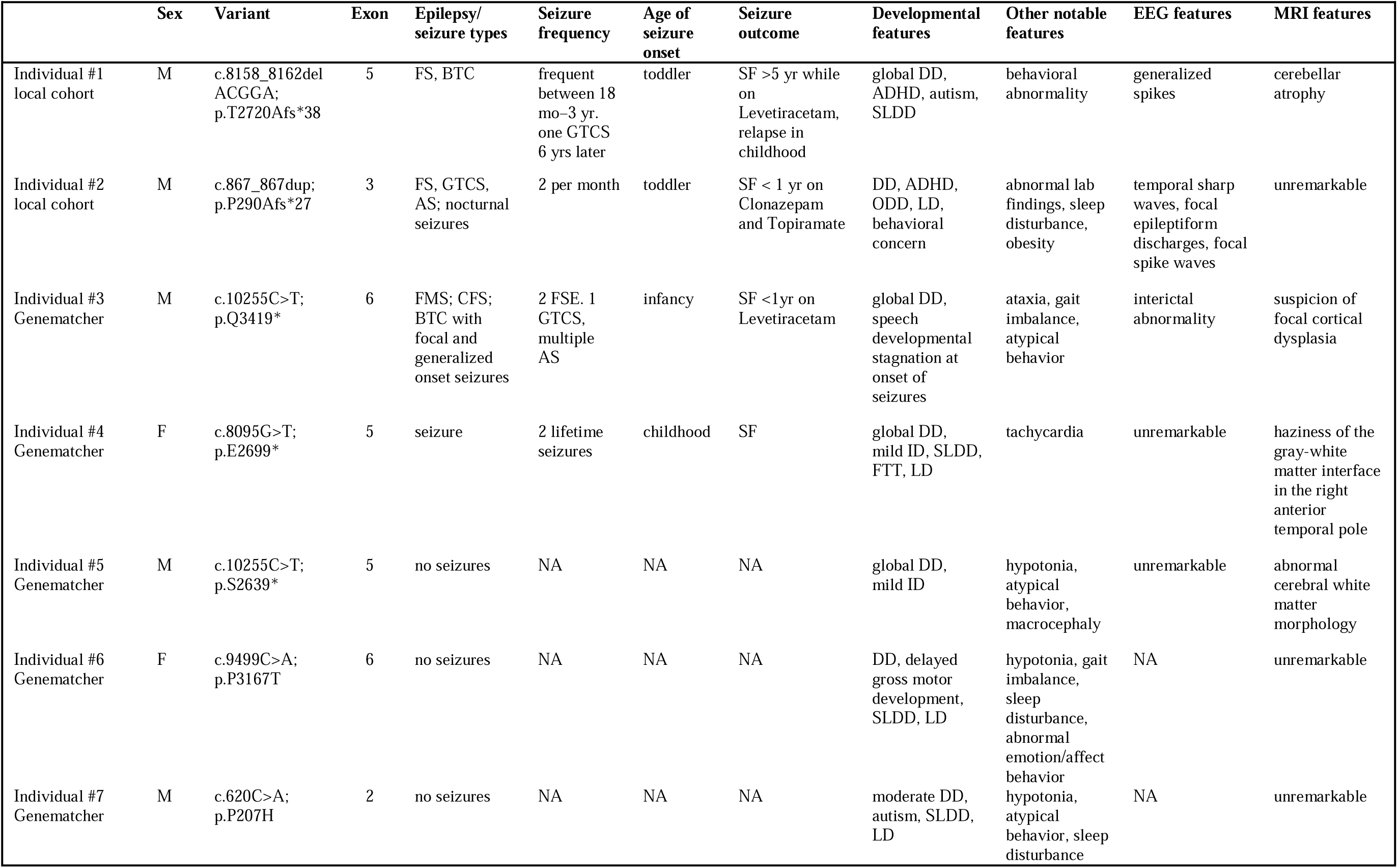
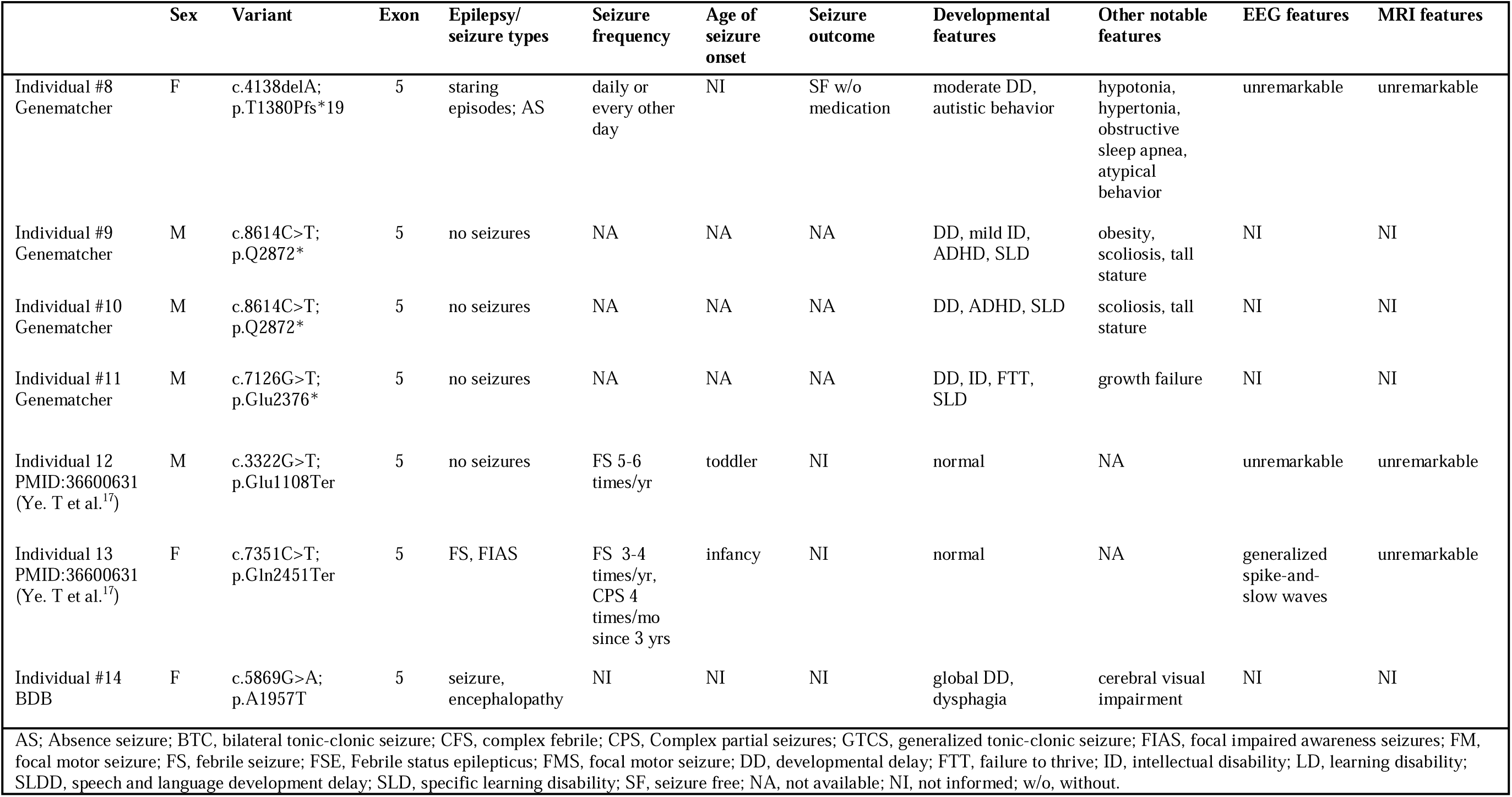
Clinical and genetic features in 14 individuals with *de novo BSN*-related disorders.

### Individuals with overlapping neurodevelopmental features carry *de novo* variants in *BSN*

We identified 12 additional individuals with confirmed *de novo BSN* variants absent from gnomAD: nine individuals through a collaborative network facilitated through Genematcher, one individual through a local biobank (BDB), and two individuals previously reported in the literature (**Table 1**).^17,24,25^ The specific variants in these 12 individuals included nine PTVs and three missense variants, which were distributed across all the functional domains of the BSN protein (**Figure 1**). Ten of the 12 variants were located in exon 5 of *BSN*; however, this likely reflects the size of this exon rather than a true mutational hotspot.

We identified overlapping seizure and developmental features in individuals with *de novo BSN* variants, consistent with those observed in both initial participants (**Table 1**). Clinically, 9/12 individuals presented with developmental delays, with 6/12 showing behavioral features such as ADHD (n=3), autistic behavior (n=2), and learning disabilities (n=6). Epilepsy was observed in half (6/12), with a median seizure onset of 16 months (range: 1–8 years). Seizure types varied; three had febrile seizures at onset, and two progressed to bilateral tonic-clonic seizures. Two individuals had epileptic encephalopathy or atypical absence seizures.

At the most recent clinical follow-up, five individuals had achieved seizure freedom for at least a year, with a median duration of 6 years (range: 1–10 years). Seizure freedom was typically achieved by a median age of 4.2 years (range: 1–4 years). Responses to treatment varied among participants. While some individuals achieved seizure freedom with anti-seizure medications such as levetiracetam, clonazepam, carbamazepine, or a combination of these, other individuals benefitted from adjunctive strategies like a ketogenic diet. Three individuals continued to have active epilepsy during the study period, with one individual experiencing seizure recurrence after 8 months of seizure freedom.

Among the six individuals without seizures, all had developmental delays, including mild intellectual disability (n=2) and/or learning disabilities (n=5). Individual #6 had developmental stagnation in infancy, but later regained skills. This was followed by fine motor delays, expressive language delays, behavioral outburst with anxiety, and sleep abnormalities. Individual #7, had language delays, self-injurious behavior, and sleep disturbances.

Additional clinical features associated with *de novo* variants in *BSN* included hypotonia (4/12) and growth abnormalities (3/12), encompassing both growth failure and tall stature. Four out of 12 individuals for whom brain imaging was available had non-specific findings, including mild cerebellar atrophy (Individual #1), suspicion of focal cortical dysplasia (Individual #3), haziness of the gray-white matter interface (Individual #4), and abnormal cerebral white matter morphology (Individual #5).

### Rare *BSN* PTVs show variable expressivity and incomplete penetrance

To investigate a potential gene-disease relationship, we analyzed the phenotypes of individuals with rare *BSN* variants absent from gnomAD that were identified via GeneMatcher (n = 5) and additional biobank databases (n = 10, CAG and PMBB, **Table S1**).^24,25^ In total, 15 individuals were found to have PTVs, including nine frameshift variants, five nonsense variants, and one splice-site variant (**Figure 1**). Of the 15 individuals with PTVs, 13/15 individuals had variants of unknown inheritance, while 2/15 individuals had paternally inherited variants including one frameshift variant and one nonsense variant (**Table S1**). Individual #19, who inherited a frameshift variant from their father, had multiple febrile seizures and bilateral tonic-clonic seizures, whereas the father, carrying the same variant, only had a single febrile seizure with no other neurological symptoms. Individual #18 had developmental delays, speech and language delays, and hyperactivity; this patient inherited a nonsense variant from their father, who did not have seizures or other neurological features. Inheritance of *BSN* variants in both individuals suggests both incomplete penetrance and variable expressivity.

Most PTV variant carriers had phenotypic features overlapping with those seen in individuals with *de novo BSN* variants, such as delayed speech and language development (4/15), global developmental delays (4/15), and specific learning disabilities (3/15). Seizures were present in 5/15 individuals, with febrile seizures as the initial presentation in 3/5 individuals.

Among the seven adults with *BSN* PTVs identified through biobank databases, 3/7 individuals did not have neurological phenotypes recorded in their EHRs. Of the 4/7 individuals with neurological phenotypes in the EMR, a single individual (Individual #29) had a seizure-related ICD-10-CM code documented **(Table S1**), while the remaining individuals had sleep apnea (G47.33), cerebral edema (G93.2), and abnormal movement (R25.2). This suggests that *BSN*-related phenotypes are comparatively mild in adulthood with incomplete penetrance.

### Comparative phenotyping of *BSN* variants identifies age-related differences

In our combined cohort of 29 individuals, we annotated 455 HPO terms across 15 phenotypic categories (**Table 2**, **Figure S1**, and **Table S2**), referred to as base terms. The most common base HPO terms were global developmental delay (HP:0001263; 45%), obesity (HP:0001513; 34%), specific learning disability (HP:0001328; 34%), and delayed speech and language development (HP:0000750; 27%). The median number of HPO terms assigned per individual was 13, with a range of 1–73 terms (**Table 2**). Through structured data harmonization and propagation, we derived 1,637 HPO terms across 616 distinct phenotypic categories, allowing for a comprehensive analysis of clinical manifestations associated with *BSN* variants (**Table 2** and **Figure S1**).^32^ The most common HPO terms after propagation were abnormality of mental function (HP:0001249; 69%), and neurodevelopmental abnormality (HP:0012759; 55%, **Figure 2A** and **Table S3**).

**Figure 2.**
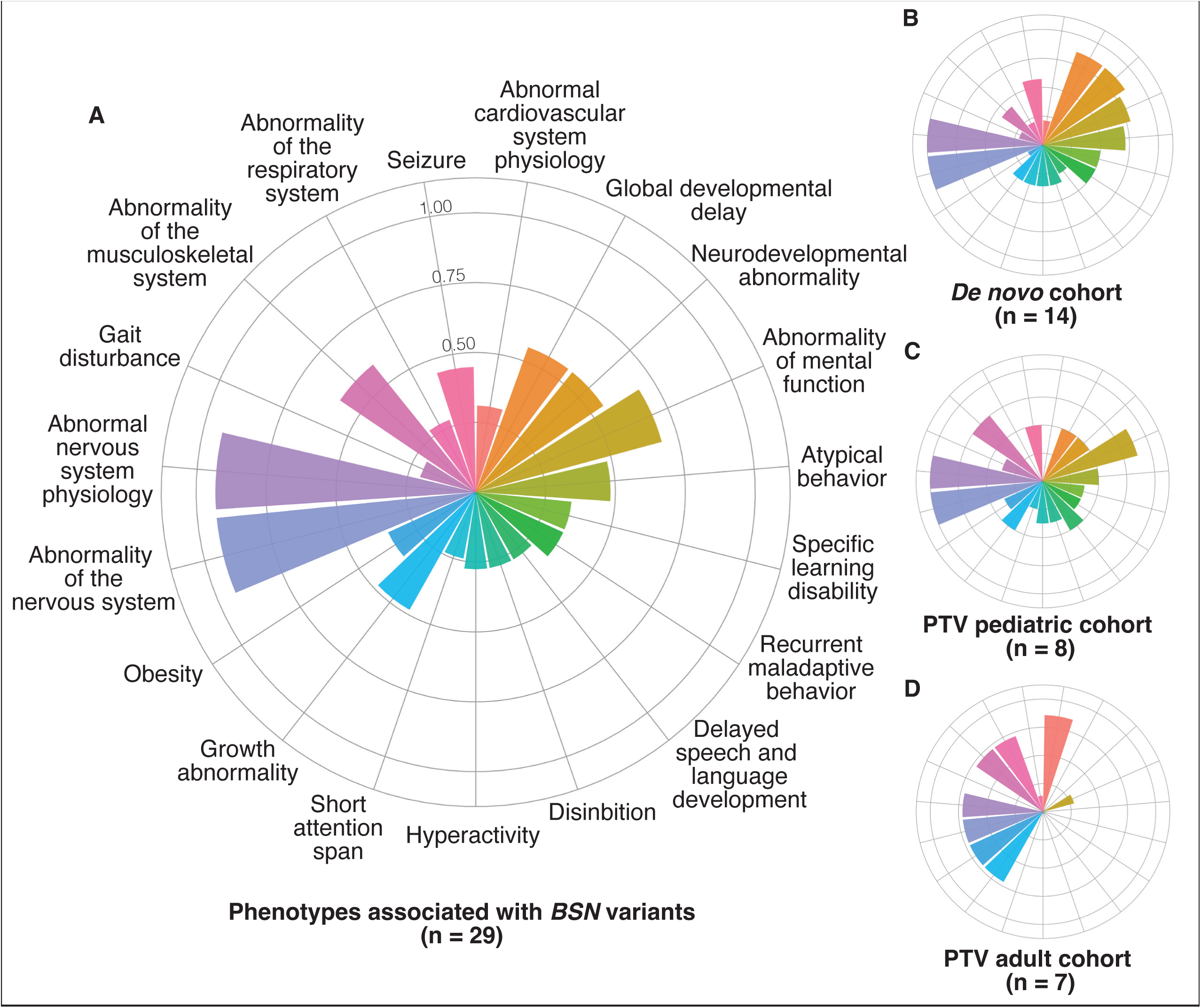
Comparison of Phenotypic Features in *BSN* Cohorts Reveals Distinct Trends by Age and Inheritance. (A) A radial plot showing phenotypic features in the overall cohort (n = 29). Radial lines reflect the frequency of specific terms within the cohort. (B) Radial graphs displaying phenotypic feature distribution across subgroups, categorized by age and inheritance: de novo cohort (n = 14, B), PTV pediatric cohort (n = 8, C), and PTV adult cohort (n = 7, D).

**Table 3.** Cohort Information on 29 individuals with rare BSN variants including de novo variants (n=14) and protein-truncating variants (n=15). Literature and biobanks report with limited data.

|  |  |
| --- | --- |
| **Demographic information** |  |
| Male | 14/29 (48.2%) |
| Female | 15/29 (51.8%) |
| **Age distribution, median (range)** |  |
| Age at assessment (*n* = 15) | 10 years (0.8 months to 85 years) |
| Seizure onset (*n* = 14) | 2.2 years (0.0 to 36 years) |
| Seizure offset (*n* = 8) | 9 months (1 month to 42 years) |
| Not applicable <sup>a</sup> | 14/29 (48.2%) |
| **Phenotypic information, median (range)** |  |
| Number of phenotypic terms per individual | 13 terms (1–73 terms) |
| Number of phenotypic terms per individual after propagation of HPO terms | 48 terms (8–224 terms) |
| Number of distinct phenotypic terms in cohort | 274 terms |
| Number of distinct phenotypic terms in cohort after propagation of HPO terms | 616 terms |

Next, we compared three groups of individuals with *BSN* variants to assess whether inheritance and age impacted phenotypic expression (**Figure 2B–2D**), including (1) a cohort of individuals with *de novo* variants of any type (*de novo* cohort, n=14), (2) individuals with PTVs of known or unknown inheritance recruited into pediatric biobanks (PTV pediatric cohort, n=8), and (3) individuals with PTVs of unknown inheritance recruited into adult biorepositories (PTV adult cohort, n=7). Both the *de novo* cohort and PTV pediatric cohorts exhibited more cognitive and seizure-related HPO terms compared to the PTV adult group. The frequency of global developmental delay (HP:0001263, p-value [p] 6.21e-08) was notably higher in those with *de novo* cohort (86%) compared to those with pediatric PTVs (50%), suggesting a potential association of developmental delays associated with *de novo BSN* variants, though recruitment bias cannot be ruled out. While both pediatric groups (*de novo* cohort and PTV pediatric cohort) displayed similar frequencies for disinhibition (HP:0000734, 36%), hyperactivity (HP:0000752, 36%), specific learning disability (HP: 0001328, 43%), and seizures (HP:000125, 57%), certain traits showed notable differences. For instance, delayed speech and language development (HP:0000750) were more prevalent in the PTV pediatric cohort (50%, p=0.004) compared to the *de novo* cohort (29%, p=0.004). Additionally, atypical behavior (HP:0000708) was observed more frequently in the *de novo* cohort (71%, p=0.004) than in those with PTVs (50%, p=0.004). Specific learning disability (HP:0001328) was also slightly more common in the *de novo* cohort (50%) than in the PTV pediatric cohort (38%), but the difference was not significant (p=0.12).

Certain seizure types were more common in the *de novo* cohort compared to the PTV pediatric cohort (**Figure 2B** and **Table S3**). Notably, focal-onset seizures (HP:0007359, 21%) and focal impaired awareness seizures (HP:0002384, 14%) were present in the *de novo* cohort but neither were reported in the PTV pediatric cohort. There were no differences between both cohorts in the frequency of febrile seizures (HP:0002373, 25%) and bilateral tonic-clonic seizures (HP:0002069, 38%).

Notably, obesity (HP: 0001513) was more common in individuals with pediatric PTVs (38%) compared to the *de novo* cohort (14%, p=0.0001). Furthermore, in the PTV adult cohort, 72% of individuals exhibited obesity-related features. Information on adults with PTVs in *BSN* was collected from PMBB, an EHR-linked biobank, which often captures more common medical conditions. Consequently, the PTV adult cohort showed a higher frequency of HPO terms related to common medical condition in adults, such as abnormal cardiovascular system physiology (HP: 0011025, 86%), a broad HPO term that includes a variety of specific clinical terms related to cardiovascular health, and abnormality of the respiratory system (HP: 0002086, 72%), which represents a high-level HPO for all medical conditions related to respiratory issues including asthma, bronchitis, and emphysema. In contrast, we did not identify a substantial frequency of high-level HPO terms indicative of neurological conditions (**Figure S1** and **Table S3**). Only 4/7 (57%) of individuals in the PTV adult cohort were found to have an abnormality of the nervous system (HP:0000707), the parent term for a wide range of neurological features, including epilepsy, movement disorders, intellectual disability, and autism.

### Association analysis reveals unique phenotypic features in *BSN*-related disorders

We reconstructed the clinical presentation of *BSN*-related disorders using 675,109 HPO terms in 14,895 probands with developmental and epileptic encephalopathies (DEEs) and neurodevelopmental disorders (NDDs) derived from various data sources including EGRP, DDD, and Epi4k (dbGaP) (**Figure 3**).^28^ To identify phenotypic features associated with *BSN*, we performed an association analysis using Fisher’s exact test comparing the frequency of HPO terms in all 29 individuals with variants in *BSN* to a larger cohort of 1,470 individuals with DEEs and 13,425 individuals with NDDs.^28^ Prominent phenotypic features associated with *BSN* variant affected individuals included disinhibition (HP:0000741, p=3.39e-17), fatigue (HP:0012378, p=5.27e-14), hypercholesterolemia (HP:0003124, p=4.01e-10), temper tantrums (HP:0025160, p=2.18e-05), and febrile seizures (HP:0002373, p=1.26e-06) as some of the most prominent phenotypes (**Figure 3** and **Table S4**). While hypercholesterolemia emerged as a notable feature in this analysis, this association may reflect the contribution of PTV adult cohort rather than a defining characteristic of *BSN*-related disorders.

**Figure 3.**
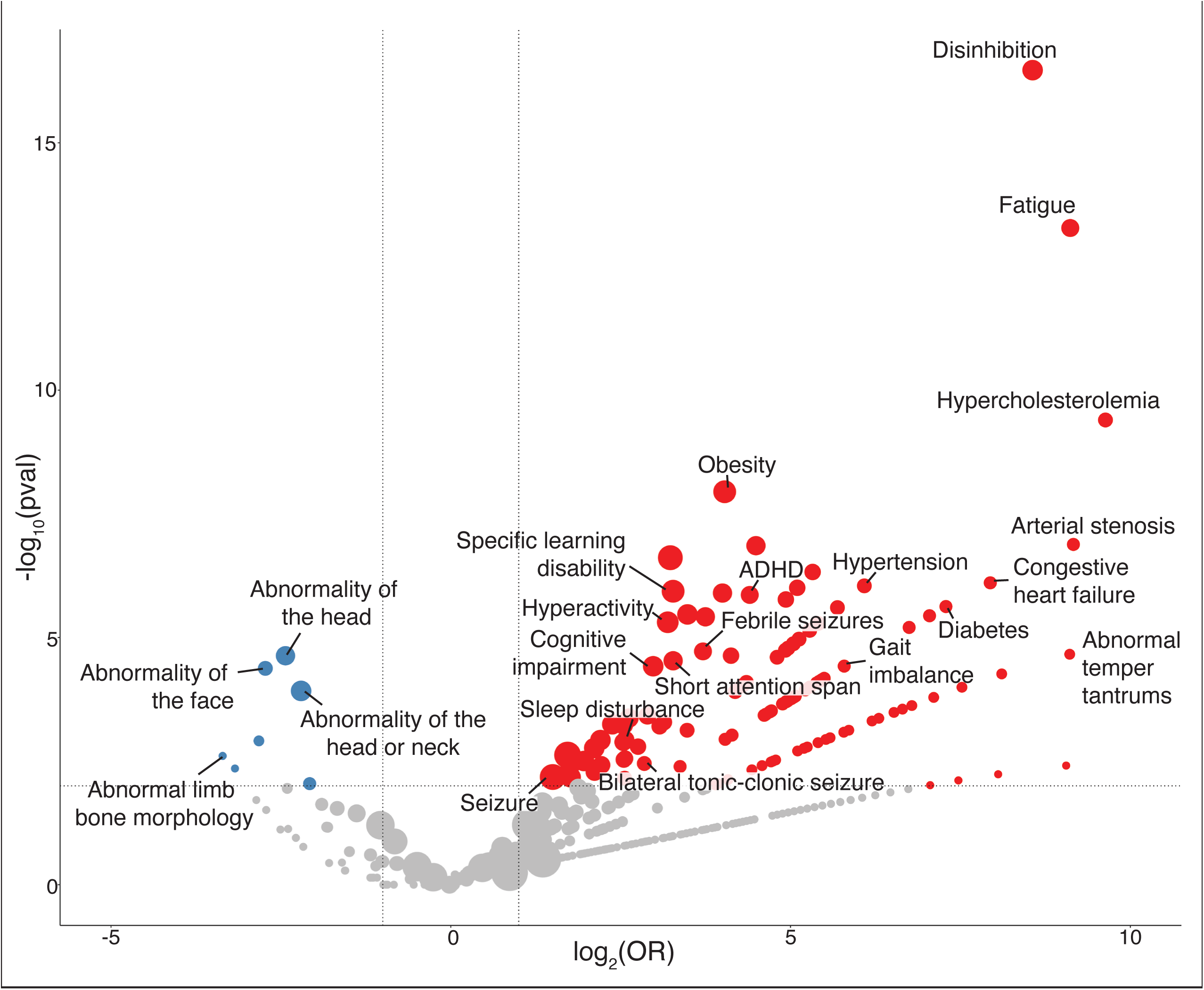
Phenotypic Association Analysis Identifies Disinhibition and Fatigue as Key Features of BSN-Related Disorders. Volcano plot depicting the frequency of HPO terms in the BSN cohort (red, n=29) compared to a larger reference group (blue, n = 14,893). Red dots represent terms with odds ratio (OR) > 0.5 and p < 0.05, indicating significant phenotypic associations in the *BSN* cohort, while blue dots represent terms with a lower association in the reference cohort. Dot size reflects term frequency within the respective group.

### Phenotypic similarity analysis supports a shared *BSN* gene-phenotype signature

Following the identification of distinct phenotypic features in individuals with BSN *variants* through association analysis, we sought to evaluate whether the phenotypic terms linked to individuals carrying *BSN* variants were sufficiently distinct to establish a discrete gene-specific phenotypic signature. We performed a formal phenotypic similarity analysis to assess the extent of clinical relatedness among individuals with *de novo BSN* variants compared to those with *de novo* variants in 256 other NDD-related genes (**Figure 4**).^28^ In brief, a phenotypic similarity analysis assesses whether clinical features observed in a subset of individuals are more related than expected by chance within a given cohort. This analysis allowed us to compare the statistical evidence for *BSN* based on phenotypic similarity with the genetic evidence derived from the relative frequency of *de novo* variants, which compares the observed versus expected number of *de novo* variants within a given gene.^31^ Individuals with *de novo BSN* variants demonstrated a significant degree of phenotypic similarity (p=0.00014), suggesting a strong gene-phenotype relationship and consistent phenotypic expression (**Figure 4**). This indicates that individuals carrying *BSN* variants had phenotypic features that are more similar than expected by chance, supporting the hypothesis of a phenotypic signature specific to *BSN*.

**Figure 4.**
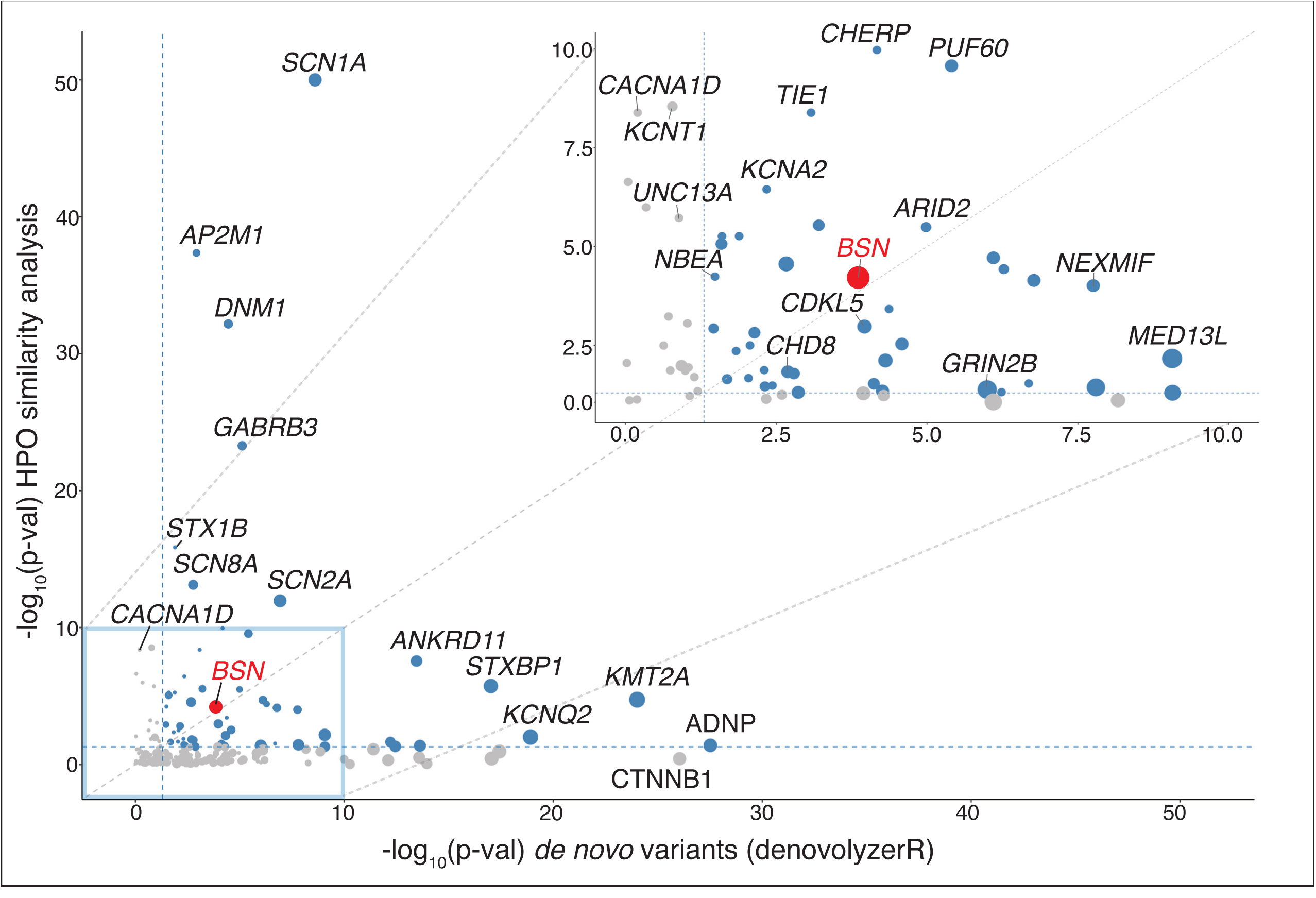
*BSN*-related Disorders Have Significant Phenotypic Resemblance in a Comparison Analysis of Genetic and Phenotypic Evidence Across 256 Genetic Etiologies Implicated in Neurodevelopmental Disorders. Scatter plot comparing genetic and phenotypic evidence for *de novo* variants in *BSN* (red, n = 14, p = 0.00014) against 257 genetic etiologies. Each data point represents an individual gene, with point size indicating the number of individuals with de novo variants per gene. Dashed blue lines denote the significance threshold of −log10(0.05) for both axes, with genes above these thresholds shown in blue to denote statistical significance in either genetic or phenotypic evidence, while genes below the thresholds are shown in gray. Genetic evidence on the x-axis reflects the statistical significance of observed de novo variants, calculated using denovolyzeR, while phenotypic evidence on the y-axis represents phenotypic similarity scores generated by sim analysis (simmax), followed by permutation analysis to assess significance. This comparative approach highlights the alignment or divergence between genetic and phenotypic evidence across genes, identifying where one type of evidence deviates from the expected correlation.

Although the phenotypic similarity for individuals carrying *de novo* variants in *BSN* was significant, the median sim score was lower compared to established genetic etiologies for epilepsy and neurodevelopmental disorder including *SCN1A* (MIM: 182389) and *SCN2A* (MIM: 182390), reflecting the variability observed in the phenotypic expression of *BSN* variants (**Figure _4_**_)._19,28

Phenotypic similarity for individuals carrying *de novo BSN* variants was lower than the phenotypic relatedness assessed by a formal phenotypic similarity analysis in 78/256 other NDD-related conditions caused by *de novo* variants (**Figure 4**). This suggests that the majority of neurodevelopmental disorder caused by *de novo* variants are more recognizable than phenotypes related to *de novo BSN* variants. In fact, the clinical relatedness of individuals carrying *de novo BSN* variants ranges only in the top 30% of all neurodevelopmental disorders assessed through this analysis.

### Longitudinal EMR data uncovers neurological and behavioral trajectories in *BSN*

In order to assess the clinical trajectory of individuals carrying *BSN* variants, we mapped available electronic medical record (EMR) data from 12 individuals across a total of 103 patient years. We included longitudinal data from five individuals in the pediatric range (birth – 18 years) and seven adults (18 years and above). This analysis allowed us to recapitulate the longitudinal disease history of *BSN*-related disorders over a median observation window of 12 years (**Figure 5**). The longitudinal analysis revealed that neurological phenotypes emerged at a median age of two years (range: 2 months–17years). Seizures were documented in three individuals, starting at a median age of one year (**Figure 5A**). Febrile seizures occurred before 18 months in two individuals, followed by other seizure types including bilateral tonic-clonic seizures, emerging between mid-childhood and early adolescence (range: 5–15 years). A single individual ascertained through the PMBB had seizures in mid-adulthood (range: 45–50 years).

**Figure 5.**
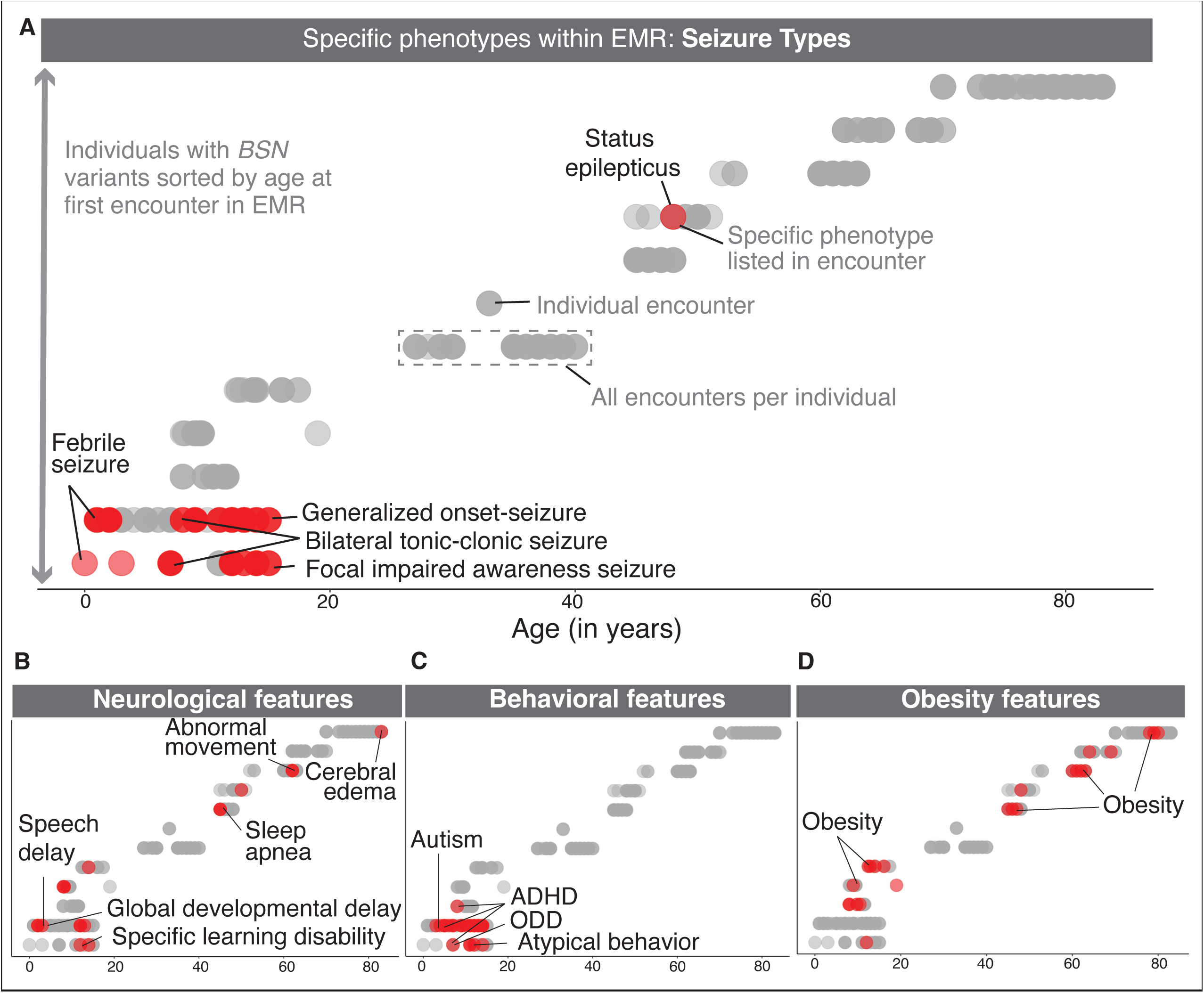
The Longitudinal Trajectory of Clinical Features in 12 Individuals with *BSN* variants Highlights Early Neurological Manifestations and Broad Phenotypic Spectrum Distribution of key clinical features (red) over time in 12 individuals with *BSN* PTVs (n=10), and *de novo* PTV variants (n=2), illustrating age-related progression of features such as epilepsy (A), neurodevelopmental delays (B), behavioral phenotypes (C), and obesity (D). Phenotypic categories were manually mapped to HPO from ICD/ICD-10 codes.

More than half of the cohort (9/12) had features of obesity, with the first recorded instance in EMR ranging widely from ten to 80 years (**Figure 5D**). Aside from the single individual with seizures recorded in the EMR in mid-adulthood (range: 45–50 years), all other adults (n=6) did not have phenotypes related to neurodevelopmental abnormalities across a cumulative time span of 61 patient years (**Figure 5B**).

Behavioral phenotypes identified through the longitudinal phenotype analysis included atypical behavior (HP:0000708) and autism spectrum disorder (HP:0000717), with ADHD (HP:0007018) diagnosed in three individuals at different ages (one in early childhood, two in late childhood) (**Figure 5C**). Neurological features were present in 4/5 individuals with longitudinal clinical data spanning infancy to adolescence (birth – 15 years), including speech delay (HP:0000750), global developmental delay (HP:0001263), and specific learning disabilities (HP:0001328). Seizures were present in 3/12 individuals with longitudinal clinical data available in the pediatric range (birth – 18 years) and between 41 to 51 years in adult individuals (**Figure 5A–B**). Obesity emerged as a common phenotype across both pediatric and adult individuals, further emphasizing the broad phenotypic spectrum associated with *BSN*-related disorders (**Figure 5D**).

Overall, our findings suggest that features associated with *BSN*-related disorders, such as febrile seizures and behavioral abnormalities, emerge during early childhood. These phenotypes diminish in frequency during adolescence and adulthood.

## Discussion

In our study, we identify *BSN*, encoding the presynaptic protein Bassoon, as a novel candidate gene for neurodevelopmental disorders and epilepsy, utilizing a multi-faceted approach that combines detailed phenotypic curation through the Human Phenotype Ontology (HPO), assessment of phenotypic similarity through computational approaches across 14,893 individuals with epilepsy and neurodevelopmental disorders, and longitudinal phenotyping analysis across 103 patient years. Phenotypic data was gathered maximizing available resources, including disease specific cohorts focused on epilepsy and neurodevelopmental disorders as well as data derived from large biobanks spanning the age spectrum. This strategy allowed us to refine the phenotypic signature of *BSN*-related variants and provide new insights into the genetic etiology across different age groups and clinical settings.^20,22,23^

We first identified two individuals with *de novo* frameshift variants in *BSN*, both of whom presented with febrile seizures early in life, which later evolved into more complex seizure types such as bilateral tonic-clonic seizures and absence seizures. Behavioral features, including ADHD and autism spectrum disorder, became evident in later childhood. Expanding our analysis, we identified 12 additional individuals with *de novo BSN* variants through collaborative efforts facilitated by GeneMatcher, biobank data, and prior literature.^17^ In total, 57% of these individuals exhibited epilepsy, with 83% experiencing febrile seizures, and several reporting multiple seizure types, such as bilateral tonic-clonic seizures (14%) and atypical absence seizures (14%).

Our findings parallel those from other genetic etiologies related to neurodevelopmental disorders, such as *SCN1A* (MIM: 182389) and *STX1B* (MIM: 601485), which exhibit a broad spectrum of seizure phenotypes, often beginning with febrile seizures in early childhood.^33,34,35^ This observation reinforces the need to consider *BSN* within the broader context of genetic etiologies related to childhood epilepsies, as our study included several individuals who transitioned from febrile to generalized seizures during adolescence (**Figure 5**). These results provide further evidence for the role of *BSN* in seizure-related neurodevelopmental disorders.

The identification of PTVs in *BSN* suggests haploinsufficiency as a likely mechanism, consistent with other genetic etiologies related to epilepsy and neurodevelopmental disorders. Insights from *Bsn* knockout mouse models strengthen this hypothesis, emphasizing the role of *BSN* in maintaining synaptic function and regulating hyperactivity of neuronal networks that may results in seizures.^14,36–38^ Homozygous *Bsn* knockout mice develop spontaneous seizures, underscoring the importance of *BSN* in regulating normal synaptic activity.^12^ Furthermore, constitutive *Bsn* mutants and GABAergic neuron-specific knockouts (*Bsn*^Dlx5^/6cKO) exhibit severe epilepsy, reinforcing the pathogenic link between Bsn disruption and epilepsy.^39^

Additionally, the presence of both missense and PTVs distributed across the gene suggests a broader disruption of protein function that may variably affect synaptic processes (**Figure 1**). Our cohort analysis, which included individuals with inherited *BSN* PTVs, provided key insights into the variability of phenotypic expression associated with this gene. Notably, 85% of the adult carriers with *BSN* PTVs were asymptomatic or only had mild neurodevelopmental phenotypes, contrasting with the more obvious presentations in pediatric individuals. This incomplete penetrance and variable expressivity have been observed in other genetic etiologies, such as *DEPDC5* (MIM: 614191), *NPRL3* (MIM: 600928), *and PRRT2* (MIM: 614386), which also show variability in clinical presentations and a relatively high proportion of asymptomatic carriers.^40–44^ Importantly, the differences observed between pediatric and adult presentations could be influenced by cohort ascertainment bias, as pediatric cohorts often focus on disease-specific phenotypes, whereas broader genetic studies include more diverse populations. By including both community-based ascertainment of individuals with *de novo* variants through GeneMatcher as well as inclusion of variant carriers in large pediatric and adult biorepositories, we believe that we have overcome such a recruitment bias and present a holistic view of the phenotypic consequences of disruptive *BSN* variants.

We utilized an HPO-based approach to obtain a larger overview of associated phenotypes in the 29 individuals carrying rare *BSN* variants harmonizing phenotypic data through the HPO ontology. This approach allowed us to identify both specific features in a relevant subset of individuals, such as febrile seizures (HP:0002373, 25%) and maladaptive behavior (HP:5200241, 35%), as well as more generalized, higher-level terms present in the majority of individuals, such as abnormality of mental function (HP:0001249, 69%) and global developmental delay (HP:0001263, 55%). The use of the HPO framework enabled us to standardize phenotypic descriptions across various cohorts, which is crucial for comparing phenotypic data in genetic studies. Our data showed that neurodevelopmental abnormalities (HP:0012759, 86%) and atypical behavior (HP: 0000708, 71%) were highly prevalent in individuals with *de novo BSN* variants. These features were also present in individuals with PTVs, although with lower frequency, highlighting the variable expressivity of *BSN* variants.

Using the same HPO-based framework to compare the 611 phenotypic features in 14 individuals with *BSN de novo* variants to 674,767 phenotypic annotations in 14,907 individuals with DEE and NDD, we identified specific features associated with *BSN*-related disorders that include disinhibition, fatigue, and febrile seizures. Furthermore, a formal phenotypic similarity analysis supported the presence of a gene-specific phenotypic signature, emphasizing that clinical features linked to disruptive *BSN* variants are more similar than expected by chance.

Identifying a gene-specific signature related to *BSN* is critical for future clinical and therapeutic studies, and this phenotypic profiling approach has provided valuable insights for genetic etiologies such as *SCN2A* (MIM: 182390) and *GRIN2A* (MIM: 138253), where a combination of *de novo* variants and phenotypic clustering has helped refine the role in neurodevelopmental disorders.^19,20,28,45^ *BSN* demonstrated moderate phenotypic similarity, suggesting that, despite phenotypic variability, many individuals carrying *de novo BSN* variants have recognizable phenotypic features. The similarity scores for *BSN*, were only higher than those seen for 179/256 other NDD-related conditions, suggesting greater phenotypic variability than 70% of all other neurodevelopmental disorder, far removed from the prominent similarity seen in *SCN1A*, *AP2M1*, or *DNM1*--related conditions.^28^ This phenotypic similarity approach provides complementary insight into gene-disease associations, supporting the link of disruptive variants in *BSN* to neurodevelopmental disorders but also quantifying the variability in phenotypic expression compared to other genetic etiologies.

Finally, our longitudinal data for 12 individuals with rare *BSN* variants illustrated that while *BSN* variants can lead to significant neurological manifestations in childhood, only a small subset of individuals had seizures and behavioral issues in adulthood (**Figure 5**). While our findings suggest that certain features, such as febrile seizures and behavioral abnormalities, tend to emerge in early childhood and may be less frequently documented in adulthood, the extent to which these features diminish over time remains uncertain. In particular, given that none of the individuals included in our study had an observation period that spanned both childhood and significant part of their adult life, it remains unclear whether the reduced frequency of neurological features in adulthood is due to the natural history of *BSN*-related disorders, or whether this observation is due to recruitment bias with more mildly affected individuals identified through large-scale biobanking in an adult cohort.

In our study, we acknowledge the uncertainty surrounding obesity as a definitive feature of *BSN*. Obesity was observed in a notable proportion of our cohort, particularly among individuals with biobank-identified PTVs (**Figure 2**). Among one of the pediatric sub-cohort (CAG) three individuals had obesity. However, these individuals were identified through a dedicated recruitment as part of an obesity research study (**Figure S2**). In contrast, only one individual in the *de novo* cohort had obesity, and the frequency of 71% in the PMBB adult cohort may reflect broader population trends rather than disease-specific associations. Prior studies have implicated *BSN* PTVs in severe adult-onset obesity, type 2 diabetes, and fatty liver disease, highlighting a potential role for *BSN* in metabolic regulation.^46,47^ Additionally, GWAS studies have linked *BSN* to both febrile seizures and obesity. Our study recapitulates these findings by identifying febrile seizures as a significant phenotypic feature and noting the occurrence of obesity in subsets of our cohort.^48^ These observations suggest a complex relationship between *BSN* variants and metabolic as well as neurodevelopmental phenotypes. Further studies are needed to determine whether these associations reflect direct effects of *BSN* disruption or cohort-specific biases, highlighting the need of integrating diverse datasets to better define the phenotypic spectrum and broader effects of *BSN* variants.

Our analysis highlights phenotypic patterns shaped by the various cohorts examined in our study, underscoring how cohort selection can influence the clinical features reported in genetic studies. By harmonizing data across multiple datasets, we strengthen our understanding of *BSN* and illustrate the importance of utilizing a wide range of study cohorts in genetic research. This unique approach enabled us to identify milder presentations of the condition that might otherwise go undetected, further highlighting the significance of integrating findings across varied populations.

A notable challenge in interpreting our findings is the distinct phenotypic presentations observed between adult and pediatric cohorts. This heterogeneity raises concerns about potential confounding factors. However, we would like to emphasize that the observed differences between cohorts actually provide important insights into the variable penetrance and expressivity of disruptive *BSN* variants. The variability in presentation suggests that our study outlines the extreme phenotypic presentations of rare *BSN* variants, ranging from unaffected adults to severe neurodevelopmental disorders with early-onset epilepsy. This variability of clinical presentations depending on patient recruitment and study cohort corroborates findings in other genetic etiologies, such as those involving *NPRL3*, *DEPDC5*, *PRRT2*, and *KCNQ2*, which highlight the challenges of delineating the full phenotypic range in variably penetrant genes.^40,41,43,44,49^

In summary, our findings position *BSN* as a novel candidate gene for neurodevelopmental disorders, demonstrating the critical interplay between genetic variants and their phenotypic manifestations. The wide range of clinical features associated with *BSN* variants delineate a new class of synaptic disorder that contrast the relative homogenous condition of other genetic etiologies linked to presynaptic function. These findings provide insight into the pathophysiology of neurodevelopmental disorders and underscore the necessity for in-depth phenotypic studies to inform relevant outcomes in gene-specific therapeutic strategies.

## Data Availability

Primary data for this analysis is available in the Supplemental material. Computer code for all analysis is available at https://github.com/staguzman/BSN/

## Acknowledgements

We thank the following who made this study possible: the biobank participants from CAG and PMBB, BDB, the research and clinical teams, and Genematcher. We acknowledge the Penn Medicine BioBank (PMBB) for providing data and thank the patient-participants of Penn Medicine who consented to participate in this research program. We would also like to thank the Penn Medicine BioBank team and Regeneron Genetics Center for providing genetic variant data for analysis. The PMBB is approved under IRB protocol (#813913) and supported by Perelman School of Medicine at University of Pennsylvania, a gift from the Smilow family, and the National Center for Advancing Translational Sciences of the National Institutes of Health under CTSA award number UL1TR001878. The CHOP Birth Defects Biorepository (BDB) is supported by the National Center for Advancing Translational Sciences, National Institutes of Health, through Grant UL1TR001878. Individual 3 was identified as part of the Acute Care Genomics study research study, funded by the Australian Government’s Medical Research Future Fund [grant number GHFM76747]. We acknowledge the use of data from the Deciphering Developmental Disorders (DDD) project. The DDD study presents independent research commissioned by the Health Innovation Challenge Fund [grant number HICF-1009-003]. This study makes use of DECIPHER, which is funded by Wellcome [grant number WT223718/Z/21/Z]. See Nature PMID: 25533962 for full acknowledgement. Figure 1 and Figure S1 were created with BioRender. Helbig, I. (2025) https://BioRender.com/n50t398.

## Web resources

DECIPHER http://www.deciphergenomics.org

GeneDx ClinVar submission page: http://www.ncbi.nlm.nih.gov/clinvar/submitters/26957/

## Contributor Information

SGG, SMR, and IH contributed to the conceptualization of the study. Data curation was performed by SGG and SMR. SGG and SG conducted the analysis, while SGG and SG developed the methods. The original draft was written by SGG, SG, SMR, and IH, and all authors (SGG, SG, SMR, and IH) participated in reviewing and editing the manuscript.

## Penn Medicine BioBank

Daniel J. Rader, M.D., Marylyn D. Ritchie, Ph.D., JoEllen Weaver, M.P.H., Giorgio Sirugo, M.D., P.h.D., Afiya Poindexter, Yi-An Ko, Ph.D., Kyle P. Nerz, Meghan Livingstone, Fred Vadivieso, Stephanie DerOhannessian, Teo Tran, Julia Stephanowski, Salma Santos, Ned Haubein, P.h.D., Joseph Dunn, Anurag Verma, Ph.D., Colleen Morse Kripke, M.S. DPT, MSA, Marjorie Risman, M.S., Renae Judy, B.S., Colin Wollack, M.S., Shefali S. Verma, Ph.D., Scott Damrauer, M.D., Yuki Bradford, M.S., Scott Dudek, M.S., Theodore Drivas, M.D., Ph.D.

## CHOP Birth Defects Biorepository

Stacy Woyciechowski, M.S., J. William Gaynor, M.D., Janine McNelia, Monica Molina, M.S., Teran Oung, Erika Diaz, M.S., Donna Stephan, M.D.

## Abbreviations

DEE: developmental and epileptic encephalopathy

HPO: Human Phenotype Ontology

NDD: neurodevelopmental disorders

