## Supplementary material for "Variants in *BSN*, encoding the presynaptic protein Bassoon, result in a novel neurodevelopmental disorder with a broad phenotypic range": Figure_S1

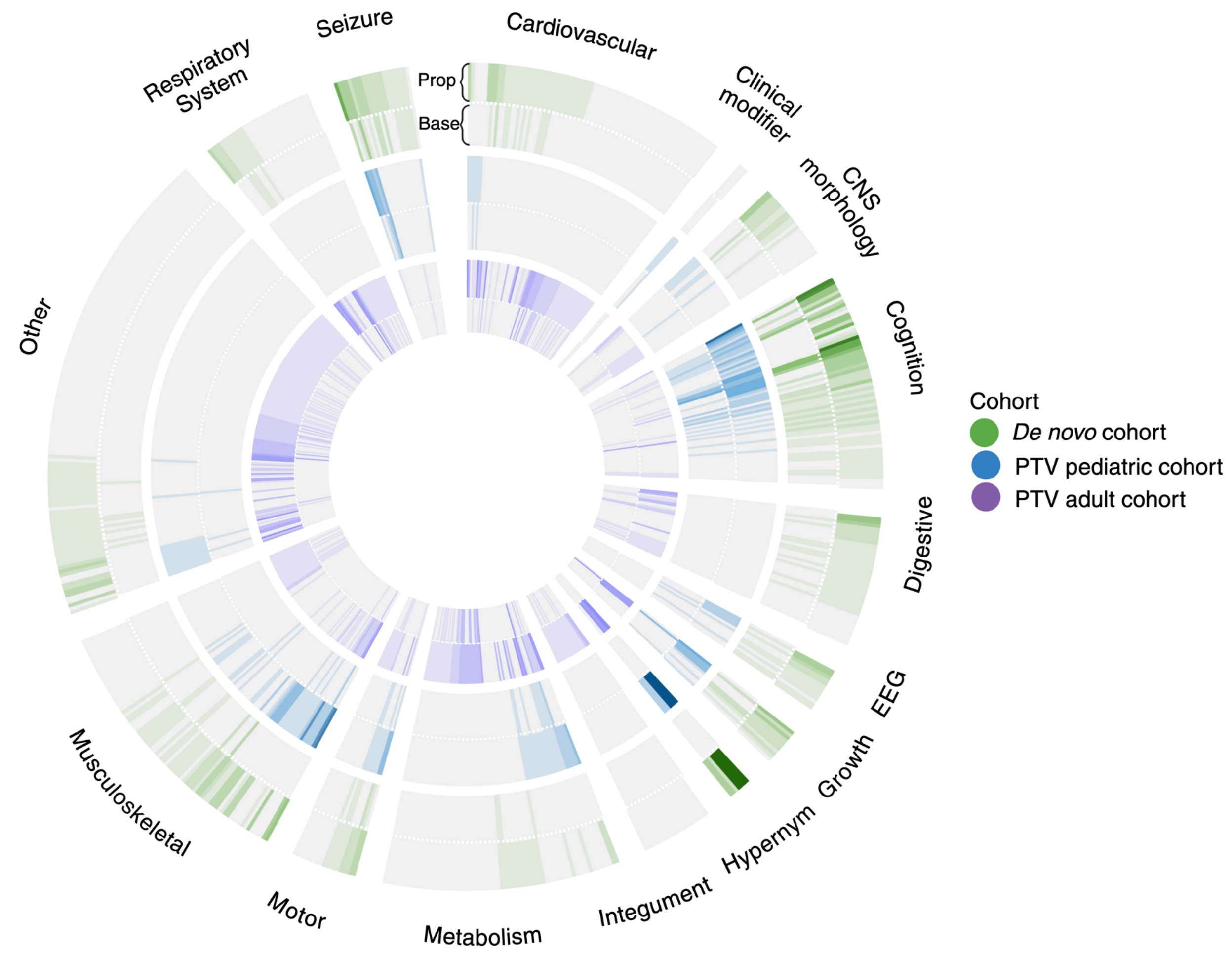


**Figure S1. Circular Plot of Observed Phenotypes in the Combined BSN Cohort, Categorized by Age, Inheritance, and Term Type**

Circular plot illustrating 616 distinct HPO terms documented in the combined cohort of 29 individuals with BSN-related disorders. Phenotypes are grouped into categories, with increased shading intensity reflecting the frequency of observed terms across individuals in the respective cohort. The outermost circle (green) represents the de novo variant cohort (n = 14), the middle circle (blue) depicts the PTV pediatric cohort (n = 8), and the innermost circle (purple) corresponds to the PTV adult cohort (n = 7). Each circle is divided into halves, with the top displaying propagated terms and the bottom showing base terms. This visualization highlights the distribution and intensity of phenotypic features across subgroups.
