## Supplementary material for "Variants in *BSN*, encoding the presynaptic protein Bassoon, result in a novel neurodevelopmental disorder with a broad phenotypic range": Figure_S2

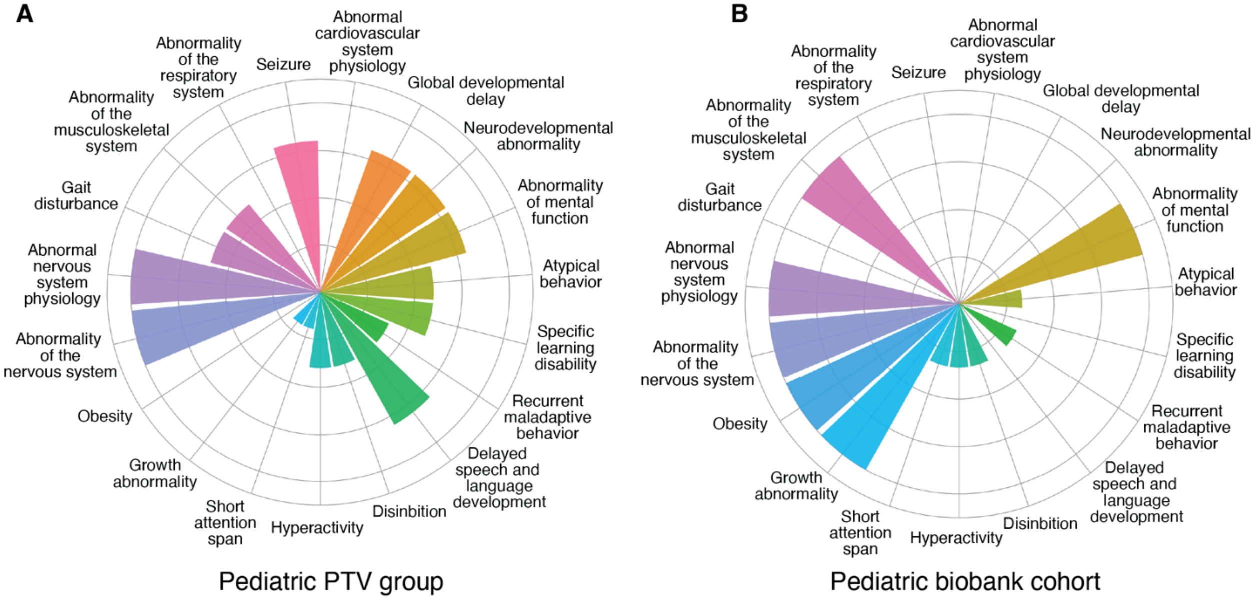


**Figure S2. Phenotypic features of *BSN* in childhood comparing individuals with PTVs and individuals within a pediatric biobank**

Radial plots illustrating the phenotypic features in the Pediatric PTV group (n = 5, A), sourced from Genematcher, and the CAG cohort (n = 3, B). Radial lines represent the frequency of specific HPO terms within each cohort. The plots highlight the distribution of phenotypic features, with separate visualizations for the Pediatric PTV and CAG cohorts to allow direct comparison.
