## Supplementary material for "Variants in *BSN*, encoding the presynaptic protein Bassoon, result in a novel neurodevelopmental disorder with a broad phenotypic range": Table_S1

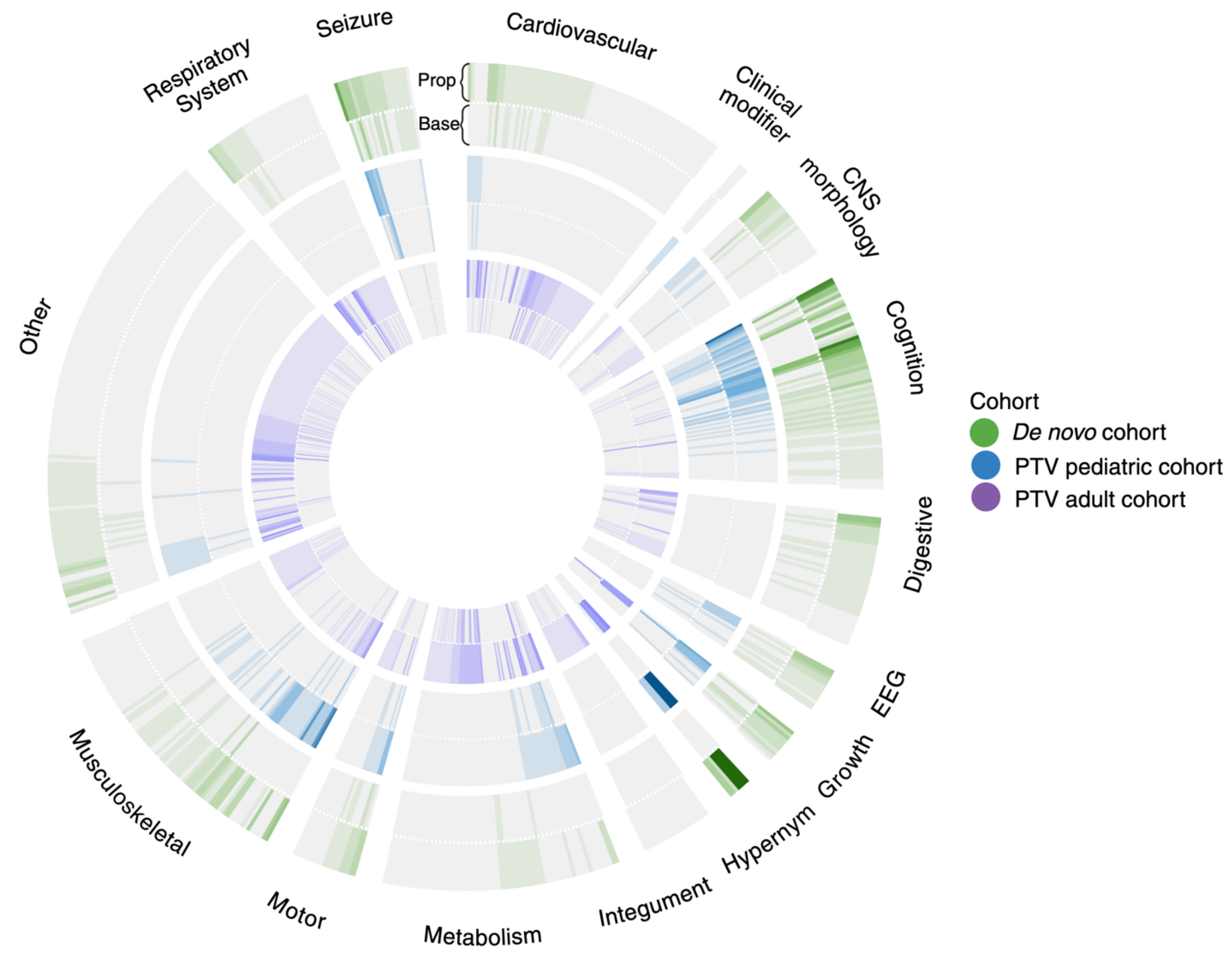


**Figure S1. Circular Plot of Observed Phenotypes in the Combined BSN Cohort, Categorized by Age, Inheritance, and Term Type**

Circular plot illustrating 616 distinct HPO terms documented in the combined cohort of 29 individuals with BSN-related disorders. Phenotypes are grouped into categories, with increased shading intensity reflecting the frequency of observed terms across individuals in the respective cohort. The outermost circle (green) represents the de novo variant cohort (n = 14), the middle circle (blue) depicts the PTV pediatric cohort (n = 8), and the innermost circle (purple) corresponds to the PTV adult cohort (n = 7). Each circle is divided into halves, with the top displaying propagated terms and the bottom showing base terms. This visualization highlights the distribution and intensity of phenotypic features across subgroups.

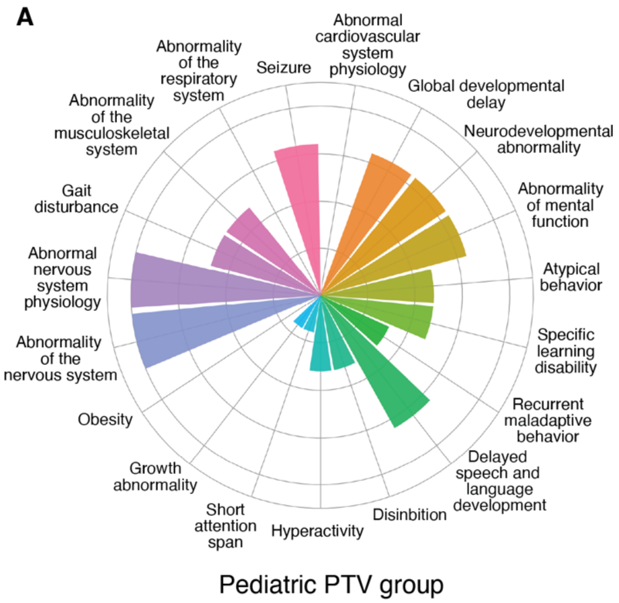


**Figure S2. Phenotypic features of *BSN* in childhood comparing individuals with PTVs and individuals within a pediatric biobank**

Radial plots illustrating the phenotypic features in the Pediatric PTV group (n = 5, A), sourced from Genematcher, and the CAG cohort (n = 3, B). Radial lines represent the frequency of specific HPO terms within each cohort. The plots highlight the distribution of phenotypic features, with separate visualizations for the Pediatric PTV and CAG cohorts to allow direct comparison.

| Table S1. Clinical and genetic features in 15 individuals with rare PTVs in *BSN*-related disorders | | | | | | | | | | | | |
|---|---|---|---|---|---|---|---|---|---|---|---|---|
| | **Sex** | **Variant** | **Exon** | **Inheritance** | **Epilepsy/ seizure types** | **Seizure frequency** | **Age of seizure onset** | **Seizure outcome** | **Developmental features** | **Other notable features** | **EEG features** | **MRI features** |
| Individual #15 Genematcher | F | c.1602delC; p.T535Pfs*7 | 4 | unknown | FS | NI | NI | NI | mild DD, mild ID, autism, ADHD, LD, SLDD | snoring issues | NI | NI |
| Individual #16 Genematcher | M | c.5840delC; p.P1947Lfs*73 | 5 | unknown | GTCS | multiple times a month first 2 yr of life | toddler | SF on Carbama zepine | mild DD, mild ID, cognitive impairment, neurodevelopm ental delay, SLDD, LD | ataxia, sleep/wake disorder, fatigue, migraine, gait imbalance, exercise intolerance | abnormally slow frequencies, focal epileptiform discharge | hyperintens ity of cerebral white matter on MRI |
| Individual #17 Genematcher | F | c.6684C>A; p.Y2228* | 5 | unknown | no seizures | NA | NA | NA | moderate DD, delayed fine motor development, autistic behavior, SLDD, LD | NI | NI | suspicion of focal cortical dysplasia |
| Individual #18 Genematcher | M | c.1027C>T; p.Gln343Ter | 3 | paternal | AS, GTCS; FS | 2 lifetime seizures | infancy | SF >2 yrs on Levetira cetam | DD, SLDD, hyperactivity | toe-walking, sound sensitivity, OSA, macrocephaly | EEG showed abnormalitie s | normal |
| Individual #19 Genematcher | M | c.3001_3006deli nsCCCTT; p.S1001Pfs*16 | 5 | paternal | FS, GTCS | 6 FS and 3 GTCS | toddler | SF >1 yr | normal | falls frequently and clumsy | normal | normal |
| Individual #20 CAG | F | c.9988C>T; p.R3330* | 6 | unknown | no seizures | NA | NA | NA | normal | Dyslipidemia, obesity | NI | NI |
| Individual #21 CAG | F | c.78delC; p.G28Afs*104 | 1 | unknown | no seizures | NA | NA | NA | normal | frequent headaches, obesity | NI | NI |
| Individual #22 CAG | F | c.9707_9708delC A; p.I3237Qfs*2 | 6 | unknown | no seizures | NA | NA | NA | ADHD | obesity | NI | NI |

Table S1. Continued

| | Sex | Variant | Exon | Inheritance | Epilepsy/ seizure types | Seizure frequency | Age of seizure onset | Seizure outcome | Developmental features | Other notable features | EEG features | MRI features |
|---|---|---|---|---|---|---|---|---|---|---|---|---|
| Individual #23 PMBB | M | c.8614C>T; p.Q2872* | 5 | unknown | no seizures | NA | NA | NA | normal | HTN, T2 diabetes, cardiac dysrhythmias, obesity | NI | NI |
| Individual #24 PMBB | M | c.8628_8629delinsA; p.L2877Wfs*17 | 5 | unknown | no seizures | NA | NA | NA | normal | heart failure, renal failure, sleep apnea, T2 diabetes, obesity | NI | NI |
| Individual #25 PMBB | M | c.7801C>T; p.R2601* | 5 | unknown | no seizures | NA | NA | NA | normal | cerebral edema, HTN, bladder cancer, obesity | NI | NI |
| Individual #26 PMBB | M | c.1986+1G>A; p.H872Pfs*10 | 4 | unknown | no seizures | NA | NA | NI | normal | ovarian dysfunction | NI | NI |
| Individual #27 PMBB | F | c.2613_2621delinsA; p.L2877Wfs*17 | 5 | unknown | no seizures | NA | NA | NI | normal | abnormal movements, GERD, HTN, T2 diabetes, obesity, | NI | NI |
| Individual #28 PMBB | F | c.2613_2621delinsA; p.L2877Wfs*17 | 5 | unknown | no seizures | NA | NI | NI | normal | breast cancer | NI | NI |
| Individual #29 PMBB | F | c.2613_2621delinsA; p.L2877Wfs*17 | 5 | unknown | status epilepticus | NI | NI | NI | normal | cardiac issues, HTN, obesity | NI | NI |

AS; Absence seizure; GTCS, generalized tonic-clonic seizure; FS, febrile seizure; DD, developmental delay; ID, intellectual disability; LD, learning disability; SLDD, speech and language development delay; HTN, hypertension; OSA, obstructive sleep apnea; SF, seizure free; NA, not available; NI, not informed; CAG, Center for Applied Genomics; PMBB, Penn Medicine BioBank.

**Table S2.** Clinical and genetic features in 15 individuals with rare PTVs in *BSN*-related disorders.
